# A Time-Dependent SEIRD Model for Forecasting the COVID-19 Transmission Dynamics

**DOI:** 10.1101/2020.05.29.20113571

**Authors:** Taarak Rapolu, Brahmani Nutakki, T. Sobha Rani, S. Durga Bhavani

## Abstract

The spread of a disease caused by a virus can happen through human to human contact or could be from the environment. A mathematical model could be used to capture the dynamics of the disease spread to estimate the infections, recoveries, and fatalities that may result from the disease. An estimation is crucial to make policy decisions and for the alerts for the medical emergencies that may arise. Many epidemiological models are being used to make such an estimation. One major factor that is important in the forecasts using the models is the dynamic nature of the disease spread. Unless we can come up with a way of estimating the parameters that guide this dynamic spread, the models may not give accurate forecasts. The main principle is to keep the model generic while making minimal assumptions. In this work, we have derived a data-driven model from SEIRD, where we attempt to forecast Infected, Recovered and Deceased rates of COVID-19 up to a week. A method of estimating the parameters of the model is also discussed thoroughly in this work. The model is tested for India at a district level along with the most affected foreign cities like Lombardia from Italy and Moscow from Russia.

The forecasts can help the governments in planning for emergencies such as ICU requirements, PPEs, hospitalizations, and so on as the infection is going to be prevalent for some time until a vaccine or cure is invented.

## 1 Introduction

Novel Coronavirus has become a pandemic within no time from the time of its detection in Wuhan, a province of China. This has been declared as a pandemic by WHO resulting in around 6,876,647 cases worldwide, by 6*^th^* of June[3]. Around 237,754 were affected in India alone. With 6,650 reported Deaths, the cases are rapidly rising, where Maharashtra is leading the tally. Its rapid progress has necessitated the need to come up with models to model the spread of the virus under different conditions like lockdown, hotspots, and migration of people across the places, and so on. The outbreak of novel coronavirus Covid19 and the ensuing utter chaos and the utter uncertainty caused by the pandemic in the entire world is unprecedented. More than ever before, it emphasizes the need for robust mathematical models that can guide policies to control the spread of infection and help in planning the hospital requirements such as PPEs, ventilators, etc [10].

In the literature several epidemiological models such as Susceptible, Infected, Recovered(SIR), Susceptible, Exposed, Infected, Recovered(SEIR) and Susceptible, Exposed, Infected, Recovered and Deceased (SEIRD), etc have been proposed to model the virus spreads like H1N1, SARS, Ebola, and others. *EpiModel* is a very useful software package, developed in ‘R’ language, that allows simulation of compartmental models, stochastic individual contact models, and the more recent network models [6].

## 2 Existing Models

A few existing epidemiological models, from which the current model is derived are discussed here. The first model used to model the pandemic virus spread is the SIR model.

### 2.1 SIR

SIR or the Susceptible, Infected, Recovered Model, this popular model [15] considers a closed population. It initially considers a small part of the population as infected. This small percentage is considered to infect *R*_0_ others, where *R*_0_ is the Basic Reproduction Rate[1]. The SIR model can be described as

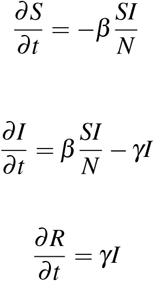

Here S, I, R stand for Susceptible, Infected and Recovered respectively. *β* is the Transmission rate and *γ* is the Recovery rate.

**Fig. 1:**
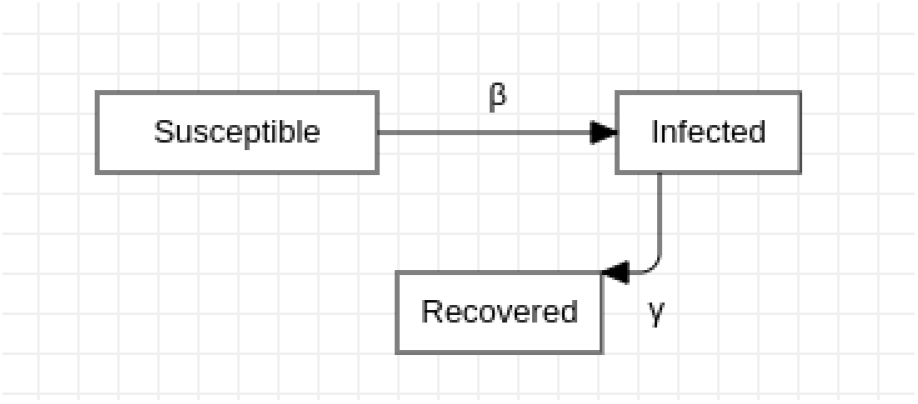
SIR Model

### 2.2 SEIR

The SIR model discussed here does not consider the percentage of the population who are exposed to the disease, but do not show any symptoms. When the incubation period i.e, the time elapsed before developing symptoms is significant, the SIR model will not be able to capture it. This leads to the SEIR model-Susceptible, Exposed, Infected, Recovered. The model is similar to SIR except that there is a transition from S to E instead of S to I. And the exposed percentage can also infect the Susceptible. In a closed population, the SEIR model can be represented as

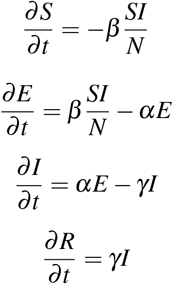

Here, *β* is the Transmission rate. *α* is the Incubation rate (Transition rate from E to I) while *γ* is the Recovery rate.

**Fig. 2:**
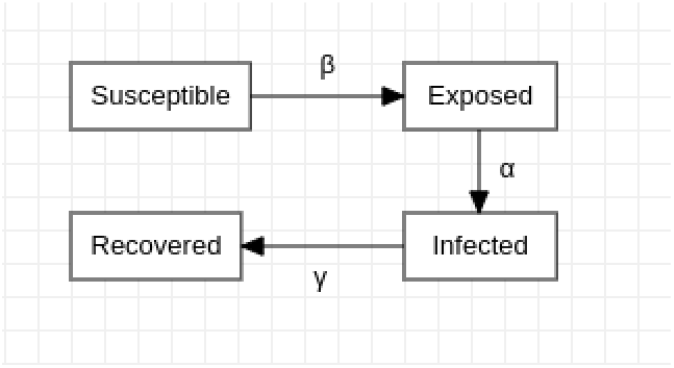
SEIR Model

Many extensions of the compartmental model have been proposed. These include extra compartments to denote, for example, the contaminated environmental reservoir [7], the eight compartment model of Tang et al.[9] to include quarantined individuals and hospitalization, etc. Most of the papers in the literature consider the SEIR model with a deterministic approach by fixing the parameters to model the spread of infection.[8][11].

Yang and Wang consider the dynamic nature of the tuning parameters themselves. They consider the time-dependent parameters to model the spread of COVID 19 virus in Wuhan extending the SEIR model [7]. They have concluded that the disease is an endemic process and requires a long term plan to spread of the virus.

The model of B. Tang et al. [4] is one of the few which considers the parameters including the rate of transmission, contact rate, recovery rate as functions of time and simulate the model in order to predict the size of the infected population. They use the Markov Chain Monte Carlo (MCMC) procedure to fit the model to the data.

We observe that one of the main challenges in adopting the compartmental models lies in tuning the number of parameters involved in the model. The work in the literature fixes the parameters based on the indicators given by epidemiological experts in the scenario. The emphasis of the current work is to estimate the parameters from the data.

## 3 Model formulation and Analysis

– Basic SEIR Model We initially ran our data against a basic SEIR model. It was observed that the results are not as accurate as expected. We were also not able to fit the Recovered rates as expected. So we extended our model to include parameter estimation-an optimized concept to estimate the parameters as per the data rather than assuming them.
– Approaches to Parameter Estimation

– Grid Search In this model, a Grid Search is used to estimate parameters. A broad range is assigned to each of the parameters. The model then tunes the parameters to get possible values that fit the data. This model is computationally expensive. It takes about an hour and a half to run it on Google Colaboratory. Once the range of parameters is narrowed down, it forecasts the rates which are more accurate than the previous model.
– Walk forward with Grid Search After working on different models, it is evident that the parameters are non-stationary i.e, they change constantly. This model implements the Walk forward approach. Until the last model, the parameters are estimated for the training set as a whole. In this model, they are estimated incrementally one day at a time. The parameters obtained for the previous day are used to estimate the current day parameters. Though this model is relatively more accurate than the previous versions, it is extremely expensive in terms of computation. Efforts are made to extend this model using Parallel computation to no avail. It took about 3-4 hours on Google Colaboratory to run this model.

## 4 Time-Dependent SEIRD Model

In order to capture the transmission dynamics, we implemented a time-dependent SEIRD model. In this model, we try to estimate parameters from the equations derived from the traditional SEIRD model rather than fitting them directly from the data. [4].

This approach resulted in forecasts of Infected, Recovered, and Deceased rates for a week and run time is also exceptionally low, compared to previous models. This model can be represented as follows:

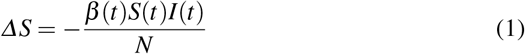

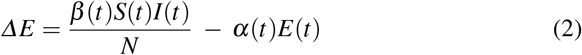

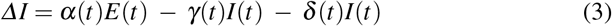

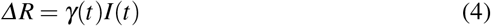

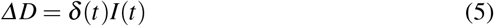

From (4), we have

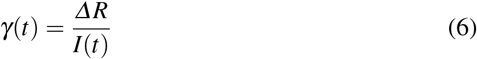

From (5), we have

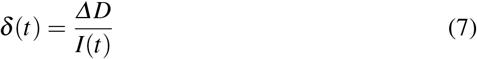

Using (6) and (7) in (3) yields

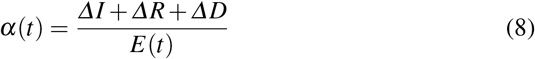

Using (8) in (2) yields

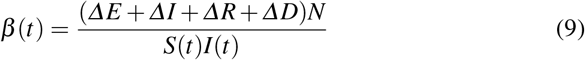

**Fig. 3:**
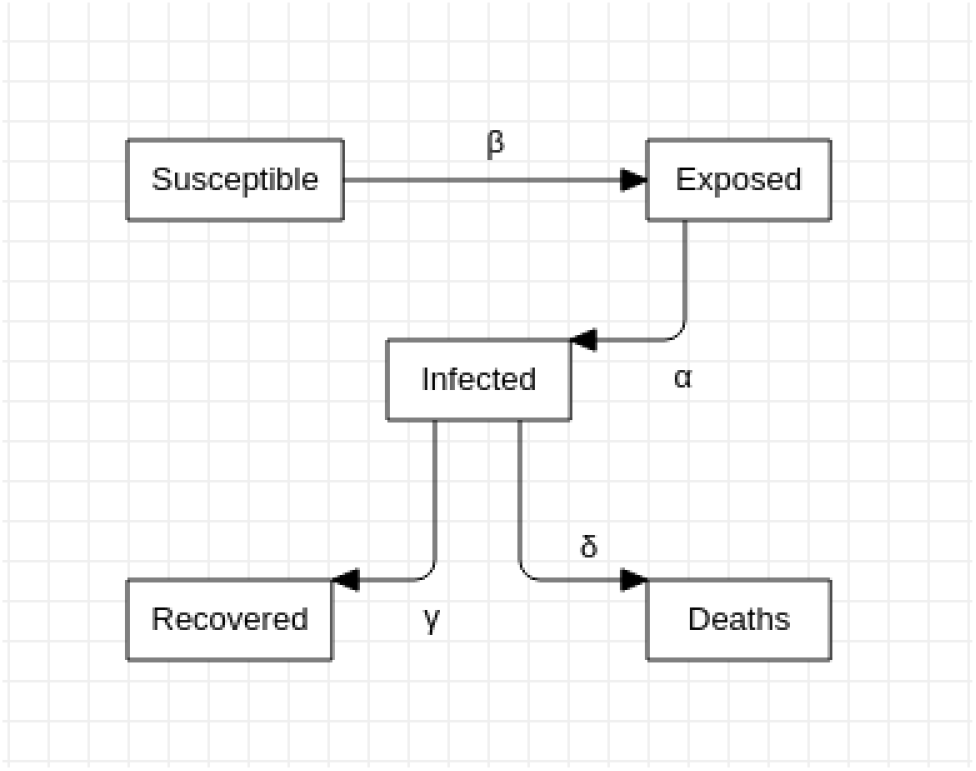
SEIRD Model

### 4.1 Data requirements and format to run the model

This model takes a *.csv* file with Cumulative Confirmed, Recovered, and Deceased values. It takes the data file, ‘N’, the population and the start-date of the lockdown of that area. Shown in Figure 4, is the format of data that is being used.

**Fig. 4:**
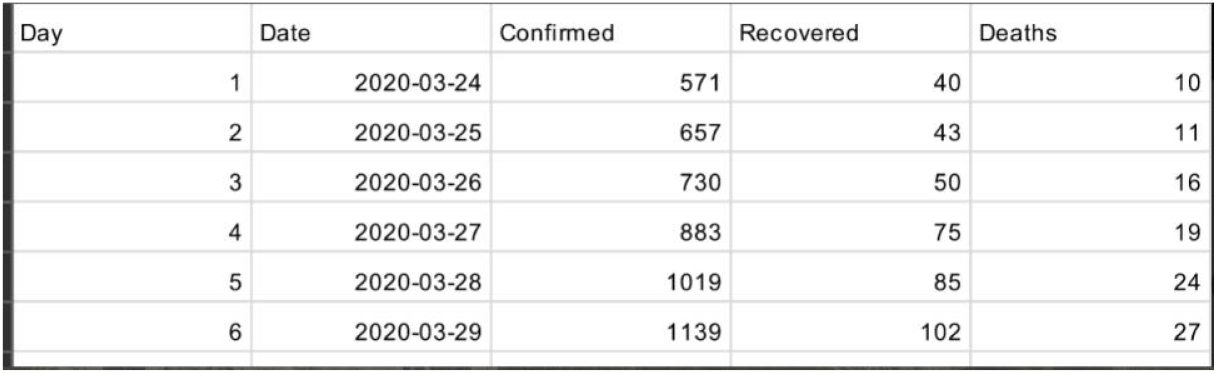
Data format

### 4.2 Model

In our model, the population is classified into 5 categories: the Susceptible, the Exposed, the Infected, the Recovered, and the Deceased. Parameters are estimated on a day to day basis using equations (6), (7), (8) and (9). The data until the previous day and the current day is used to calculate the parameters of the previous day. Then the value for the Exposed population that is calculated is passed on to calculating the parameters for the next day. It should be noted that there is a difference between the real and official data, due to the testing capability in a region. Since our model is data-driven, and we use the official data, it can forecast only the cases that will be reported

The data sources used for training the model are available at [2] for India, [14] for Moscow, Russia and [13] for Lombardia, Italy. For districts in India, the data is available from 24*^th^* April in [2]. It is stated that the Incubation Period is 2-14 days [7]. So, the forecasts after 14 days might not be accurate. Therefore we limited our prediction window to 7 days and consider the last 7 days of the data for parameter selection.

### 4.3 Parameter Selection

From the 7 days considered, we get 6 sets of model parameters (alpha, beta, gamma, delta) each. As mentioned before, we use the *D^th^* and *(D* +1 *)^th^* days’ data to calculate *D^th^* day’s parameters. Since the available deceased data is more reliable, we calculate the Mean Absolute Percentage Error or MAPE between the actual and forecasted deceased on *(D +* 2*)^th^* day and so the last day’s parameter set can not be used. As we increase the validation period, we will be losing the recent parameters. Therefore, we limited our validation period to one day. We now consider the parameter with the least deceased MAPE and use it to forecast Infected and Deceased values. If in case, two parameters have the same deceased MAPE, we select the one with the least Infected MAPE.

### 4.4 Algorithm

The code for the model is put up at the link given in [5]. Here, s[], e[], i[], r[], d[] are arrays to store calculated susceptible, exposed, infected, recovered and Deceased values

alpha[], beta[], gamma[], delta[] -arrays to store calculated alpha, beta, gamma and delta values

preds are all the predictions stored in an array pred_values are stored in a stack that contains s, e, i, r, d array values in seird function.

*start.date* is starting date of the data taken from data.csv

*T* number of days in the training data taken from data.csv

*incub-period* is the Incubation Period mape_values stores the calculated mape values.

#### Algorithm 1 Time-dependent SEIRD Model

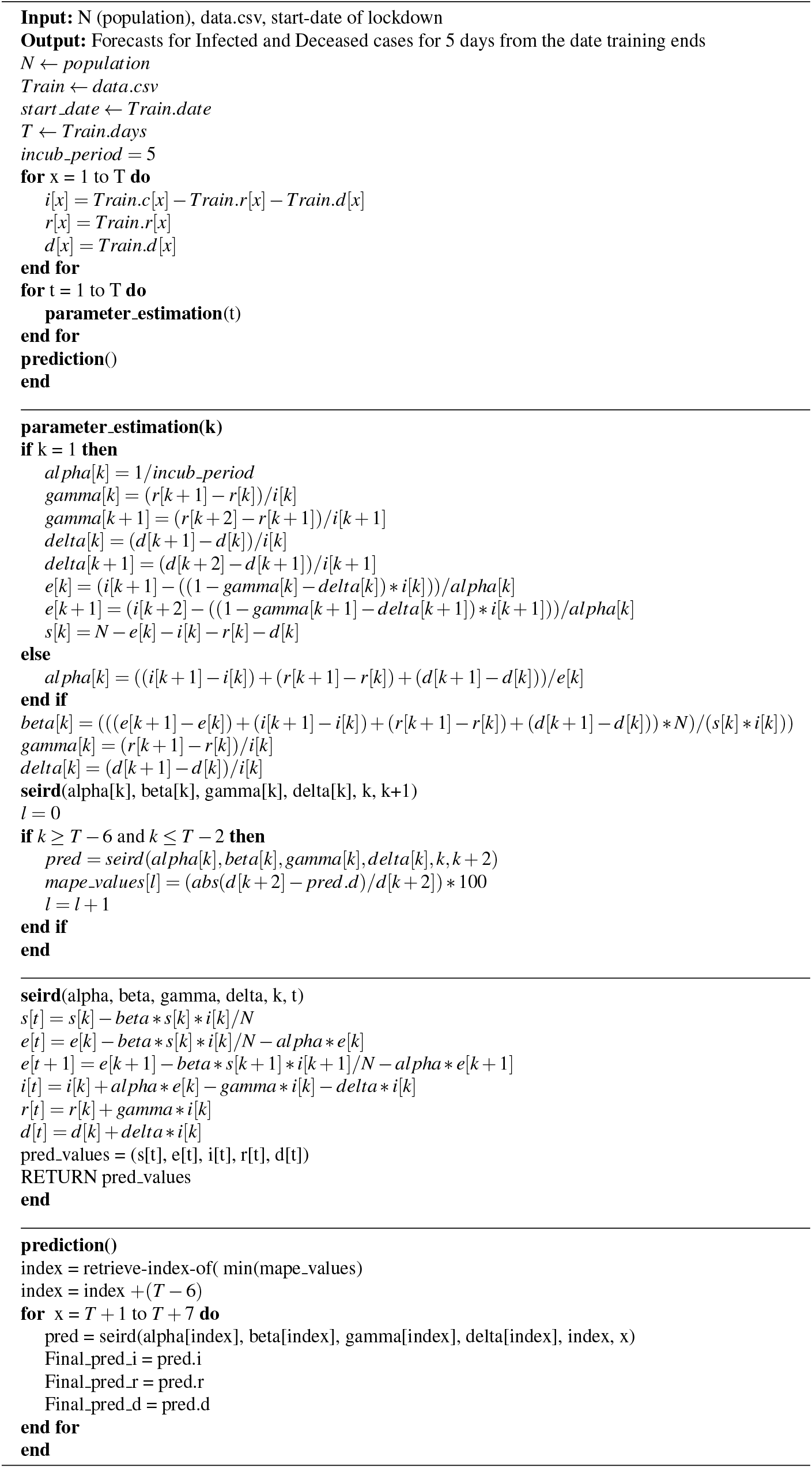

## 5 Results and Analysis

Forecasted infected and deceased plots for India 2, most affected regions of Maharashtra 4, Tamilnadu 6, Gujarat 8, Italy 12 and Russia 16 are shown in the appendix. Test data from 4*^th^* of June to 11*^th^* of June is used for the districts in India and 3*^rd^* of June to 10*^th^* June for others.

### 5.1 Analysis

Actual vs. Forecasted plots of Infected and Deceased cases for India are shown in Figure 5 and 6 repectively. For Mumbai, figures 7, 8; Chennai 9, 10; Ahmedabad 11, 12; Lombardia 13, 14 and Moscow 15, 16 are presented here for Infected and Deceased cases respectively.

**Fig. 5:**
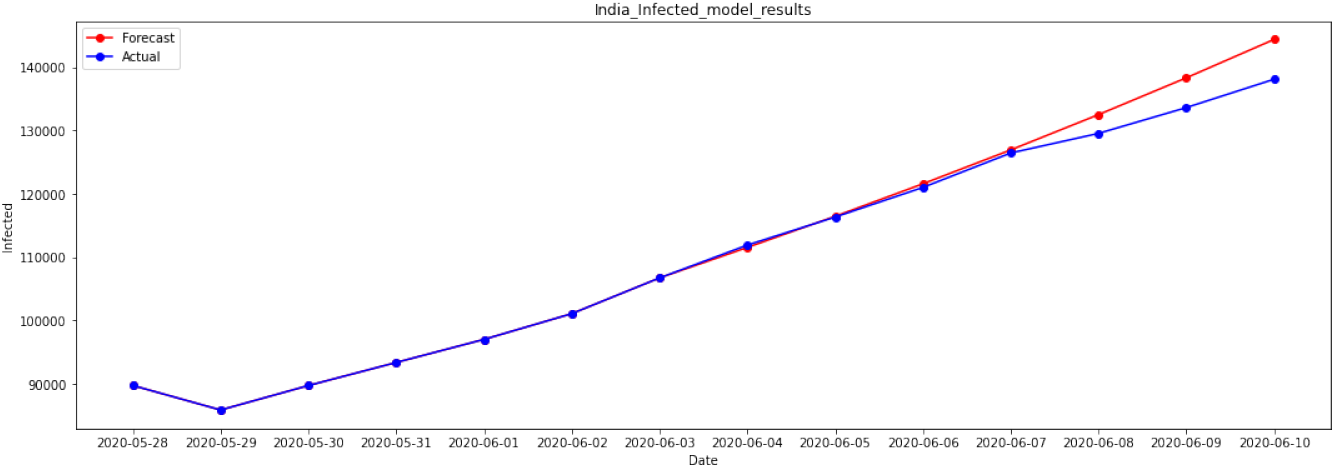
Forecasts of Infected Cases of *India* from 3*^rd^* June to 10*^th^* June

**Fig. 6:**
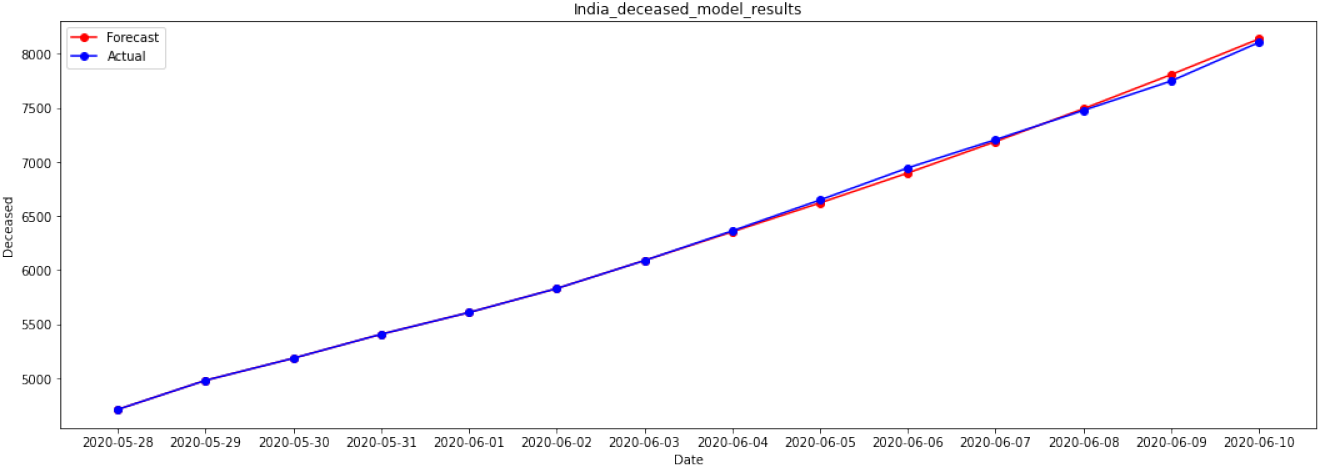
Forecasts of Deceased Cases of *India* from 3*^rd^* June to 10th June

**Fig. 7:**
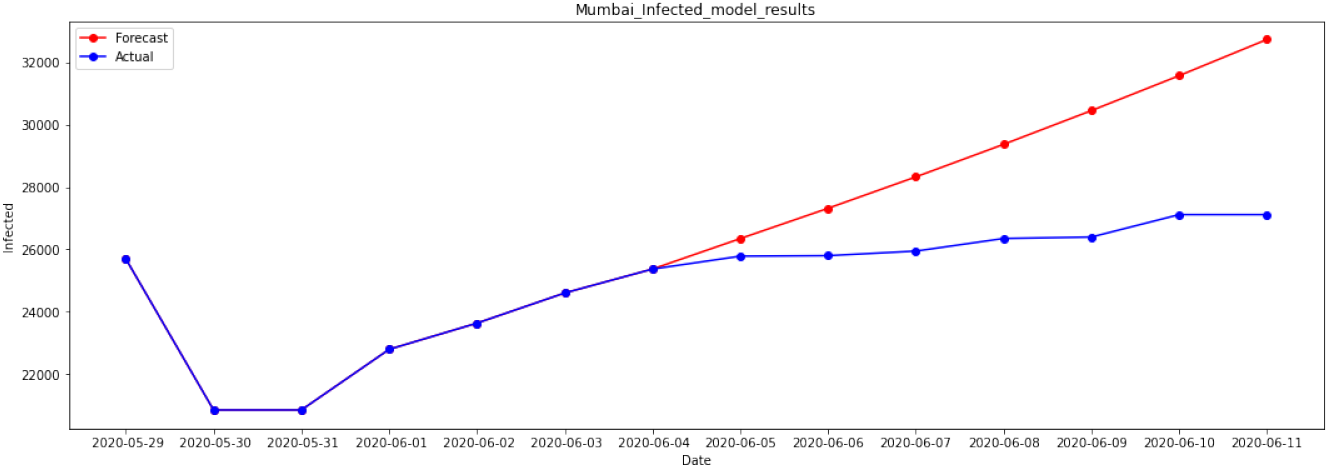
Forecasts of Infected Cases of *Mumbai* from 4*^th^* June to 11*^th^* June

**Fig. 8:**
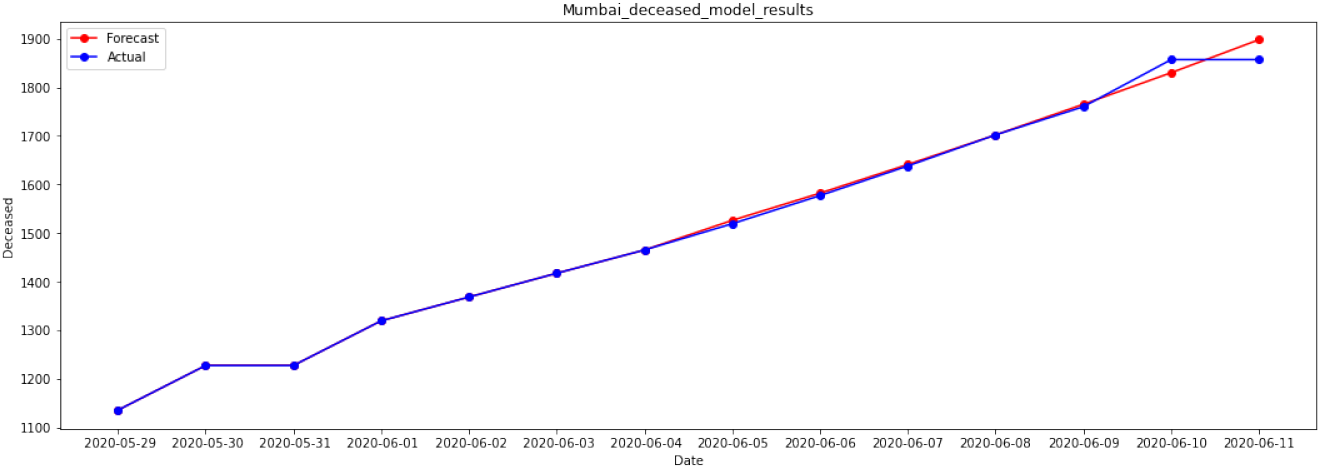
Forecasts of Deceased Cases of *Mumbai* from 4*^th^* June to 11*^th^* June

**Fig. 9:**
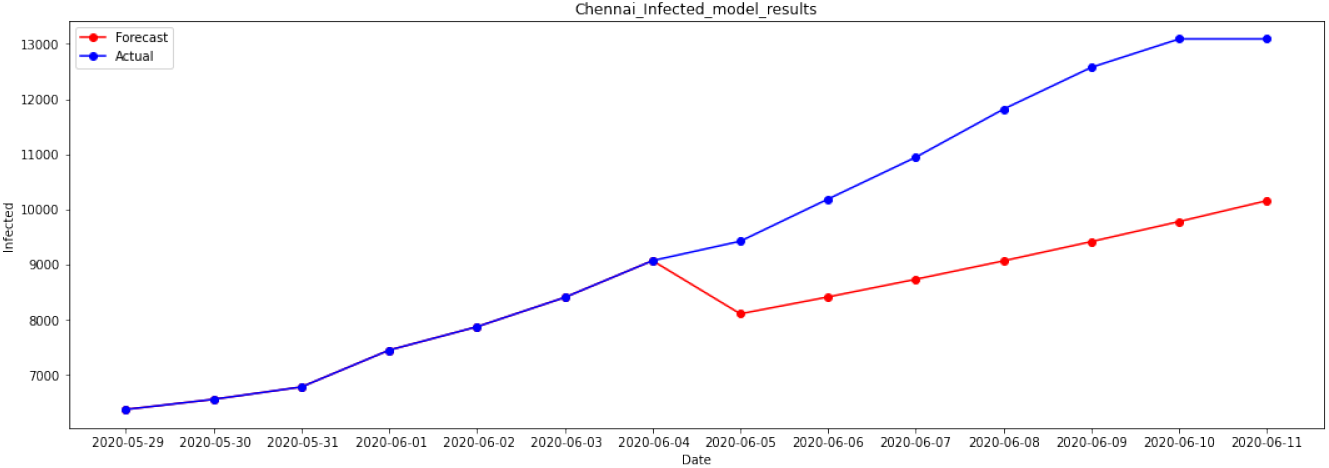
Forecasts of Infected Cases of *Chennai* from 4*^th^* June to 11*^th^* June

**Fig. 10:**
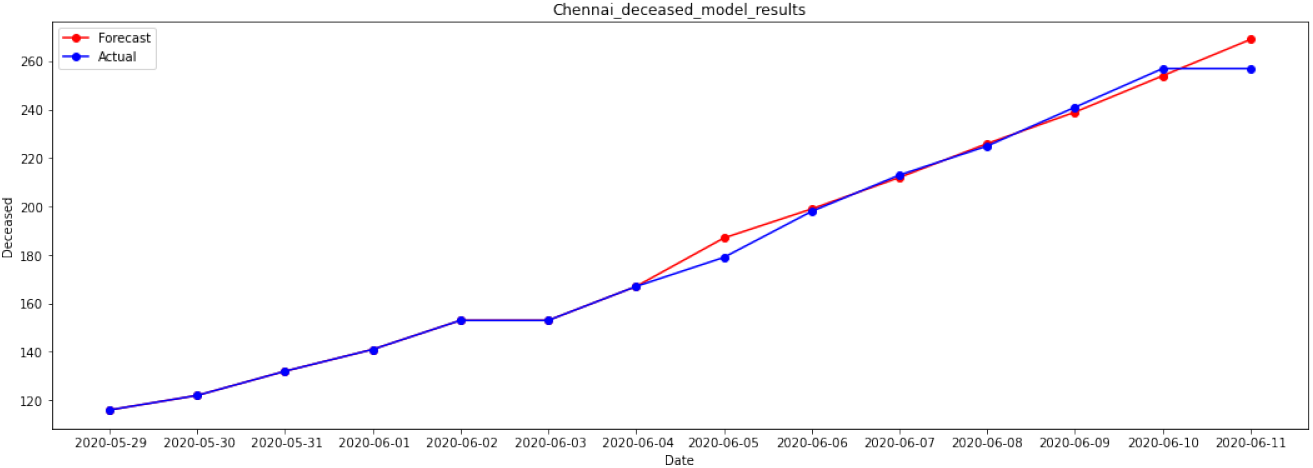
Forecasts of Deceased Cases of *Chennai* from 4*^th^* June to 11*^th^* June

**Fig. 11:**
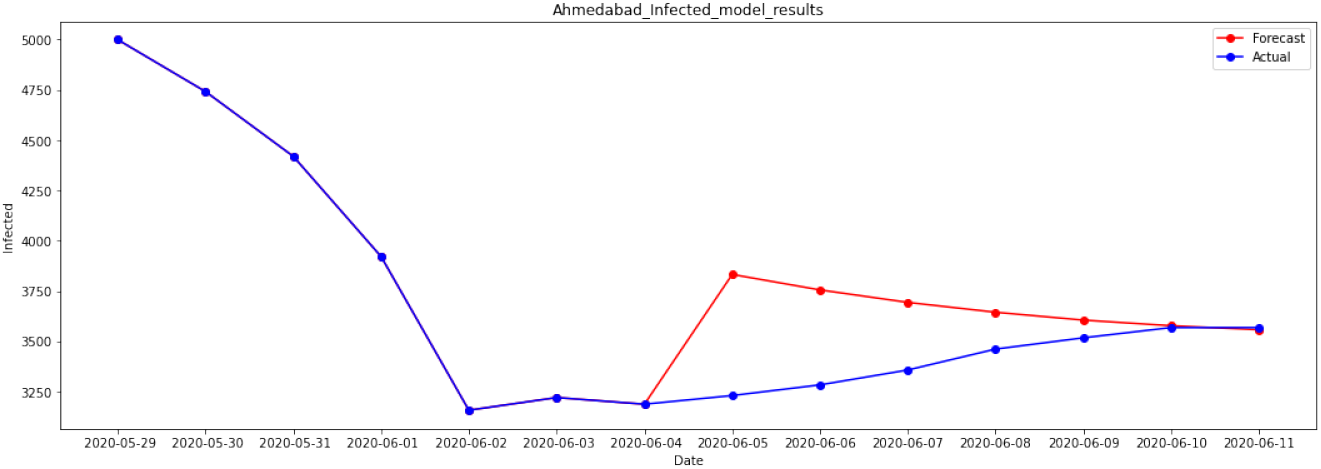
Forecasts of Infected Cases of *Ahmedabad* from 4*^th^* June to 11*^th^* June

**Fig. 12:**
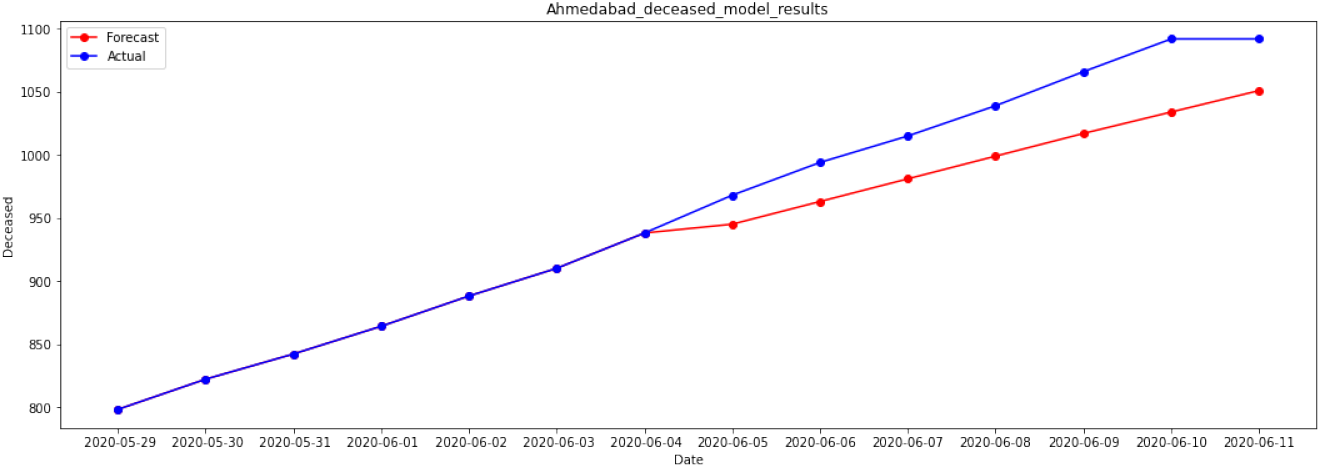
Forecasts of Deceased Cases of *Ahmedabad* from 4*^th^* June to 11*^th^* June

**Fig. 13:**
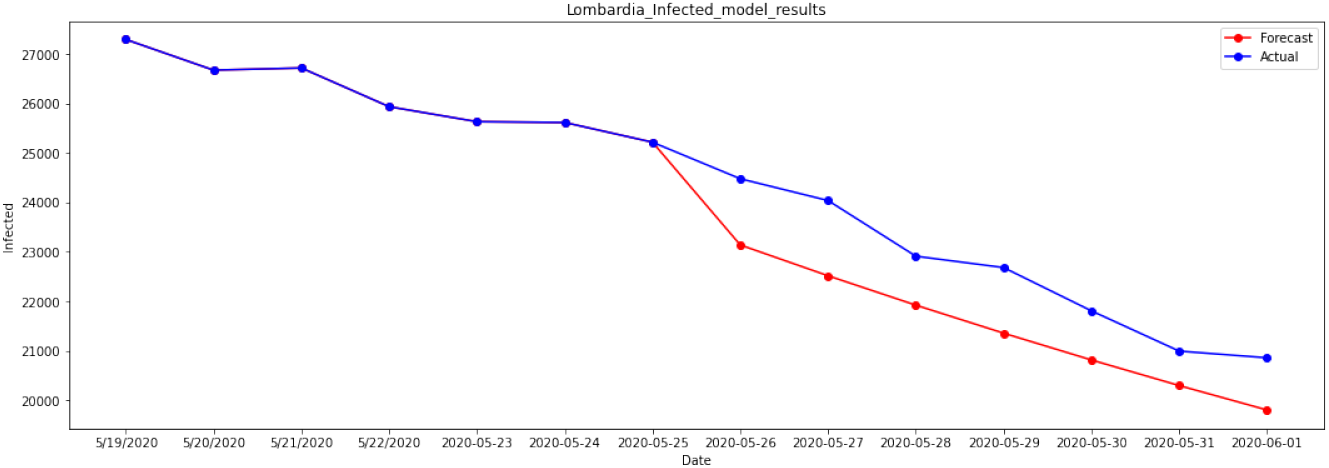
Forecasts of Infected Cases of *Lombardia, Italy* from 25*^th^* May to 1^st^ June

**Fig. 14:**
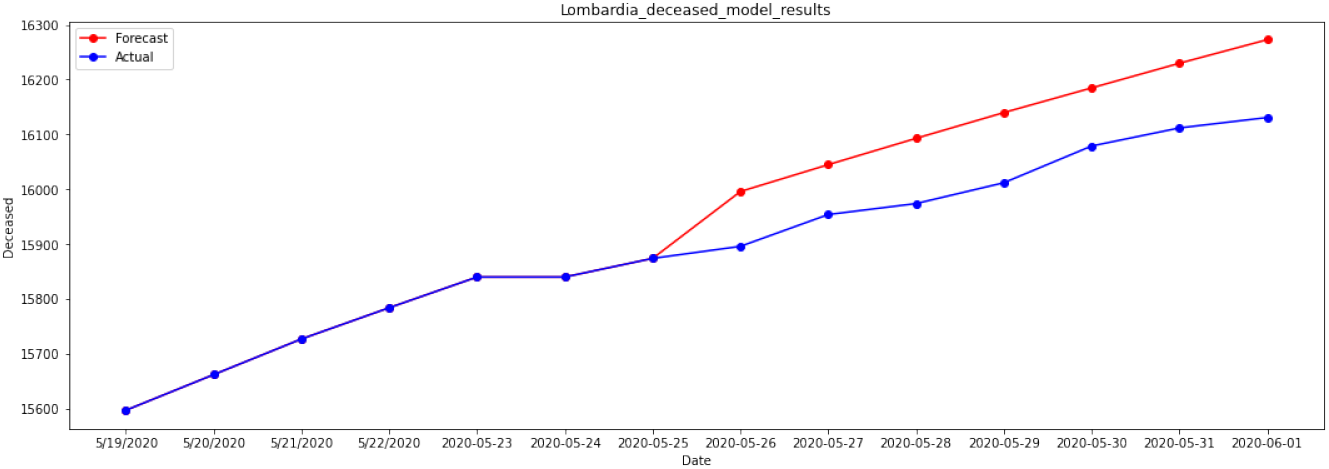
Forecasts of Deceased Cases of *Lombardia, Italy* from 25*^th^* May to 1^st^ June

**Fig. 15:**
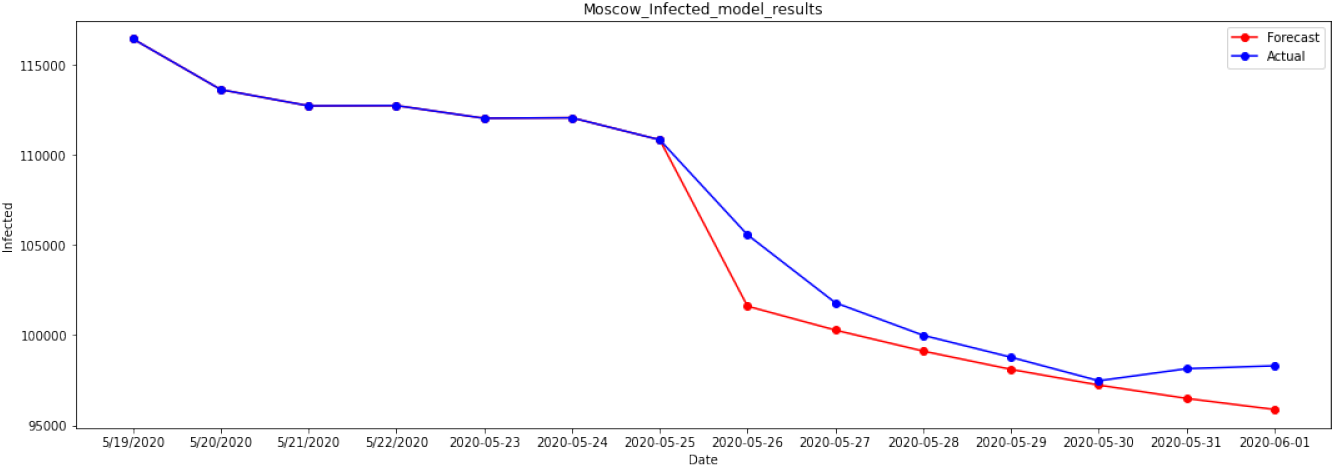
Forecasts of Infected Cases of *Moscow* from 25*^th^* May to 1*^st^* June

**Fig. 16:**
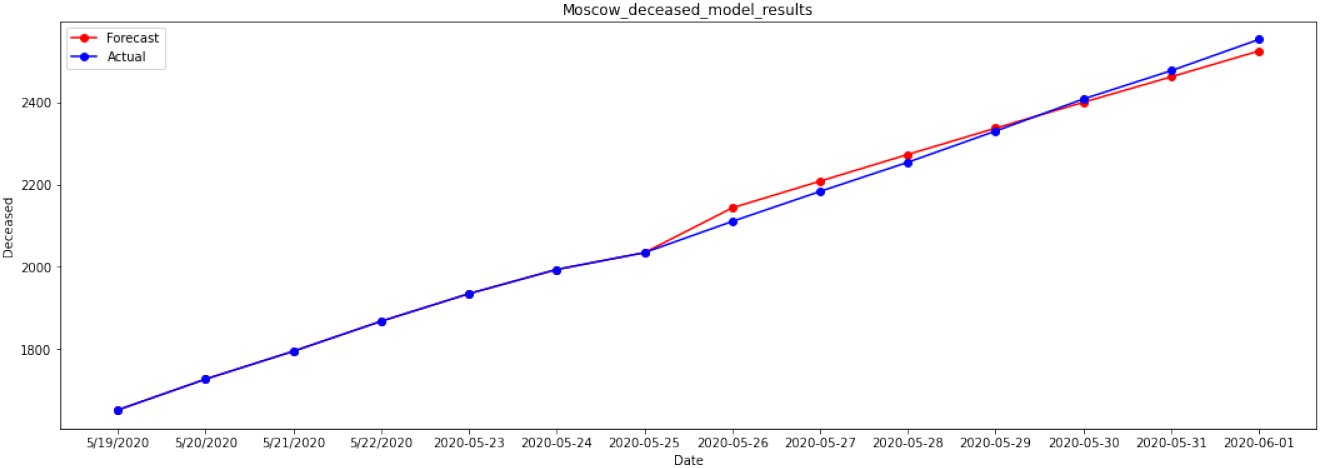
Forecasts of Deceased Cases of *Moscow* from 25*^th^* May to 1*^st^* June

From the results obtained for the above mentioned areas, it can be summarized that the model can capture the current-trend properly. Its forecasts are based on the growth rate of the actual curve. If there is a sudden increase or decrease in the growth rate, the forecasts will not be so accurate until the model stabilizes.

## 6 Extensions of the present model

As mentioned earlier, the main principles behind this model are the accuracy of the forecasts and minimum assumptions. We have taken care to avoid assumptions while building the model. As we try to add more compartments to the model, the number of parameters involved also increase. In order to estimate the parameters, either we make an educated guess or derive them from the data. This model is developed keeping India in mind. Since there is no proper data available regarding the tests, the quarantined and others for India, we did not compartmentalize our model. If any other country has the data required for compartmentalization, the model can be extended further.

## 7 Limitations

– This model considers a closed population. Birth, Mortality rates, and others are not considered.
– This model is limited to short term forecasts as the parameters keep changing and they can not be approximated to long term.
– The transmission rate for the exposed is not considered due to the uncertainty in the transmission dynamics of the exposed.

## 8 Future Scope

– Considering population density instead of a homogeneous population to forecast accurate results.
– The forecasts of Infected cases are affected by the sudden change in the data. The model has to be improved so that it need not wait until the model is stabilized.
– Transmission rate for exposed has to be tuned properly.
– Quarantine factor and others can be included to get a more detailed analysis of the situation.

## 9 Conclusions

Several papers are published using SEIR to predict the results. Initial parameters are assumed to be constant in these papers. The parameters were assumed based on input from hospitals and other sources. In this model, parameters are calculated from the data rather than making an educated guess. The goal was to forecast these results so that we can estimate and plan for Hospital equipment and Personal Protective Equipment in advance.

## Data Availability

The data referred to in this manuscript is open-sourced and is available in the below-mentioned links.

https://api.covid19india.org/

https://www.worldometers.info/coronavirus/

https://github.com/pcm-dpc/COVID-19/blob/master/dati-regioni/dpc-covid19-ita-regioni.csv

https://www.kaggle.com/kapral42/covid19-russia-regions-cases?select=covid19-russia-cases-scrf.csv

## A Tables

**Table 1:** Forecasts of Infected cases for *India*

| *S.No* | *Date* | Actual | Forecast |
| --- | --- | --- | --- |
| 0 | 2020-06-04 | $1.12 \cdot 10^5$ | $1.12 \cdot 10^5$ |
| 1 | 2020-06-05 | $1.16 \cdot 10^5$ | $1.16 \cdot 10^5$ |
| 2 | 2020-06-06 | $1.21 \cdot 10^5$ | $1.22 \cdot 10^5$ |
| 3 | 2020-06-07 | $1.26 \cdot 10^5$ | $1.27 \cdot 10^5$ |
| 4 | 2020-06-08 | $1.3 \cdot 10^5$ | $1.32 \cdot 10^5$ |
| 5 | 2020-06-09 | $1.34 \cdot 10^5$ | $1.38 \cdot 10^5$ |
| 6 | 2020-06-10 | $1.38 \cdot 10^5$ | $1.44 \cdot 10^5$ |

**Table 2:** Forecasts of Death cases for *India*

| *S.No* | *Date* | Actual | Forecast |
| --- | --- | --- | --- |
| 0 | 2020-06-04 | 6,363 | 6,355 |
| 1 | 2020-06-05 | 6,649 | 6,622 |
| 2 | 2020-06-06 | 6,946 | 6,899 |
| 3 | 2020-06-07 | 7,207 | 7,190 |
| 4 | 2020-06-08 | 7,478 | 7,493 |
| 5 | 2020-06-09 | 7,750 | 7,809 |
| 6 | 2020-06-10 | 8,107 | 8,139 |

**Table 3:**
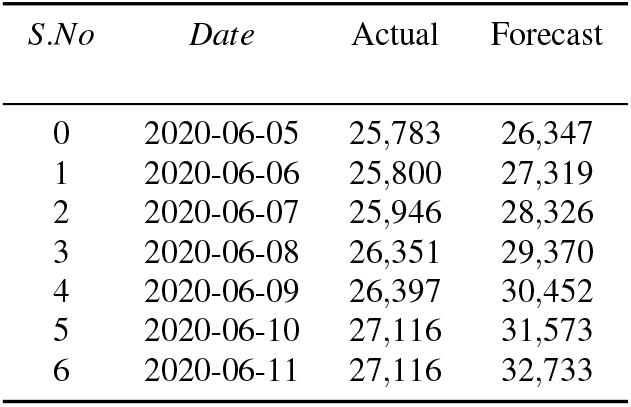
Forecasts of Infected cases for *Mumbai*

**Table 4:** Forecasts of Death cases for *Mumbai*

| *S.No* | *Date* | Actual | Forecast |
| --- | --- | --- | --- |
| 0 | 2020-06-05 | 1,519 | 1,526 |
| 1 | 2020-06-06 | 1,577 | 1,582 |
| 2 | 2020-06-07 | 1,638 | 1,641 |
| 3 | 2020-06-08 | 1,702 | 1,702 |
| 4 | 2020-06-09 | 1,760 | 1,765 |
| 5 | 2020-06-10 | 1,857 | 1,830 |
| 6 | 2020-06-11 | 1,857 | 1,898 |

**Table 5:** Forecasts of Infected cases for *Chennai*

| *S.No* | *Date* | Actual | Forecast |
| --- | --- | --- | --- |
| 0 | 2020-06-05 | 9,420 | 8,103 |
| 1 | 2020-06-06 | 10,185 | 8,409 |
| 2 | 2020-06-07 | 10,944 | 8,730 |
| 3 | 2020-06-08 | 11,817 | 9,064 |
| 4 | 2020-06-09 | 12,574 | 9,413 |
| 5 | 2020-06-10 | 13,089 | 9,777 |
| 6 | 2020-06-11 | 13,089 | 10,156 |

**Table 6:** Forecasts of Death cases for *Chennai*

| *S.No* | *Date* | Actual | Forecast |
| --- | --- | --- | --- |
| 0 | 2020-06-05 | 179 | 187 |
| 1 | 2020-06-06 | 198 | 199 |
| 2 | 2020-06-07 | 213 | 212 |
| 3 | 2020-06-08 | 225 | 226 |
| 4 | 2020-06-09 | 241 | 239 |
| 5 | 2020-06-10 | 257 | 254 |
| 6 | 2020-06-11 | 257 | 269 |

**Table 7:** Forecasts of Infected cases for *Ahmedabad*

| *S.No* | *Date* | Actual | Forecast |
| --- | --- | --- | --- |
| 0 | 2020-06-05 | 3,231 | 3,833 |
| 1 | 2020-06-06 | 3,284 | 3,756 |
| 2 | 2020-06-07 | 3,358 | 3,694 |
| 3 | 2020-06-08 | 3,462 | 3,645 |
| 4 | 2020-06-09 | 3,518 | 3,606 |
| 5 | 2020-06-10 | 3,569 | 3,578 |
| 6 | 2020-06-11 | 3,569 | 3,558 |

**Table 8:** Forecasts of Death cases for *Ahmedabad*

| *S.No* | *Date* | Actual | Forecast |
| --- | --- | --- | --- |
| 0 | 2020-06-05 | 968 | 945 |
| 1 | 2020-06-06 | 994 | 963 |
| 2 | 2020-06-07 | 1,015 | 981 |
| 3 | 2020-06-08 | 1,039 | 999 |
| 4 | 2020-06-09 | 1,066 | 1,017 |
| 5 | 2020-06-10 | 1,092 | 1,034 |
| 6 | 2020-06-11 | 1,092 | 1,051 |

**Table 9:** Forecasts from 1*^st^* June of Infected cases for *Lombardia, Italy*

| *S.No* | *Date* | Actual | Forecast |
| --- | --- | --- | --- |
| 0 | 2020-05-26 | 24,477 | 23,140 |
| 1 | 2020-05-27 | 24,037 | 22,518 |
| 2 | 2020-05-28 | 22,913 | 21,924 |
| 3 | 2020-05-29 | 22,683 | 21,358 |
| 4 | 2020-05-30 | 21,809 | 20,817 |
| 5 | 2020-05-31 | 20,996 | 20,300 |
| 6 | 2020-06-01 | 20,861 | 19,807 |

**Table 10:** Forecasts from 1*^st^* June of Death cases for *Lombardia, Italy*

| *S.No* | *Date* | *Actual* | *Forecast* |
| --- | --- | --- | --- |
| 0 | 2020-05-26 | 15,896 | 15,996 |
| 1 | 2020-05-27 | 15,954 | 16,045 |
| 2 | 2020-05-28 | 15,974 | 16,093 |
| 3 | 2020-05-29 | 16,012 | 16,140 |
| 4 | 2020-05-30 | 16,079 | 16,185 |
| 5 | 2020-05-31 | 16,112 | 16,230 |
| 6 | 2020-06-01 | 16,131 | 16,273 |

**Table 11:** Forecasts of Infected cases for *Lombardia, Italy*

| *S.No* | *Date* | *Actual* | *Forecast* |
| --- | --- | --- | --- |
| 0 | 2020-06-04 | 20,224 | 20,465 |
| 1 | 2020-06-05 | 19,853 | 20,336 |
| 2 | 2020-06-06 | 19,499 | 20,209 |
| 3 | 2020-06-07 | 19,420 | 20,084 |
| 4 | 2020-06-08 | 19,319 | 19,959 |
| 5 | 2020-06-09 | 18,297 | 19,837 |
| 6 | 2020-06-10 | 17,857 | 19,716 |

**Table 12:** Forecasts of Death cases for *Lombardia, Itlay*

| *S.No* | *Date* | *Actual* | *Forecast* |
| --- | --- | --- | --- |
| 0 | 2020-06-04 | 16,201 | 16,187 |
| 1 | 2020-06-05 | 16,222 | 16,205 |
| 2 | 2020-06-06 | 16,249 | 16,224 |
| 3 | 2020-06-07 | 16,270 | 16,242 |
| 4 | 2020-06-08 | 16,302 | 16,260 |
| 5 | 2020-06-09 | 16,317 | 16,278 |
| 6 | 2020-06-10 | 16,349 | 16,296 |

**Table 13:** Forecasts from 1*^st^* June of Infected cases for *Moscow, Russia*

| *S.No* | *Date* | Actual | Forecast |
| --- | --- | --- | --- |
| 0 | 2020-05-26 | $1.06 \cdot 10^5$ | $1.02 \cdot 10^5$ |
| 1 | 2020-05-27 | $1.02 \cdot 10^5$ | $1 \cdot 10^5$ |
| 2 | 2020-05-28 | 99,992 | 99,115 |
| 3 | 2020-05-29 | 98,774 | 98,099 |
| 4 | 2020-05-30 | 97,464 | 97,226 |
| 5 | 2020-05-31 | 98,135 | 96,485 |
| 6 | 2020-06-01 | 98,296 | 95,870 |

**Table 14:** Forecasts from 1*^st^* June of Death cases for *Moscow, Russia*

| *S.No* | *Date* | Actual | Forecast |
| --- | --- | --- | --- |
| 0 | 2020-05-26 | 2,110 | 2,143 |
| 1 | 2020-05-27 | 2,183 | 2,208 |
| 2 | 2020-05-28 | 2,254 | 2,273 |
| 3 | 2020-05-29 | 2,330 | 2,337 |
| 4 | 2020-05-30 | 2,408 | 2,400 |
| 5 | 2020-05-31 | 2,477 | 2,462 |
| 6 | 2020-06-01 | 2,553 | 2,525 |

**Table 15:** Forecasts of Infected cases for *Moscow, Russia*

| *S.No* | *Date* | Actual | Forecast |
| --- | --- | --- | --- |
| 0 | 2020-06-04 | 91,750 | 93,454 |
| 1 | 2020-06-05 | 90,905 | 92,857 |
| 2 | 2020-06-06 | 89,766 | 92,333 |
| 3 | 2020-06-07 | 89,384 | 91,877 |
| 4 | 2020-06-08 | 89,701 | 91,488 |
| 5 | 2020-06-09 | 85,824 | 91,160 |
| 6 | 2020-06-10 | 83,167 | 90,893 |

**Table 16:** Forecasts of Death Cases for *Moscow, Russia*

| *S.No* | *Date* | Actual | Forecast |
| --- | --- | --- | --- |
| 0 | 2020-06-04 | 2,749 | 2,769 |
| 1 | 2020-06-05 | 2,806 | 2,840 |
| 2 | 2020-06-06 | 2,864 | 2,910 |
| 3 | 2020-06-07 | 2,919 | 2,980 |
| 4 | 2020-06-08 | 2,970 | 3,050 |
| 5 | 2020-06-09 | 3,029 | 3,120 |
| 6 | 2020-06-10 | 3,085 | 3,189 |

